# Sibling Psychiatric Disorders and Risk of Postpartum Psychosis

**DOI:** 10.1101/2025.04.22.25326225

**Authors:** Shelby Smout, Floriana Milazzo, Thalia K. Robakis, Veerle Bergink, Behrang Mahjani

## Abstract

**Objective:** Postpartum psychosis affects 1-2 per 1,000 women following childbirth. Although bipolar disorder and prior postpartum psychosis are established risk factors, the contribution of familial psychiatric history is not well characterized.

**Methods:** Using Swedish national registers, we identified 1,213,829 women with a full sibling who delivered their first liveborn child between 1980 and 2017. Postpartum psychosis was defined as mania, psychosis, or psychotic depression within 90 days postpartum. Associations between sibling psychiatric diagnoses and postpartum psychosis were estimated using logistic regression with cluster-robust standard errors, including stratification by the woman’s own psychiatric history.

**Outcomes:** Overall, 1,831 women developed postpartum psychosis (1.5 per 1,000). In sister-sister pairs, the strongest associations were for sister’s postpartum psychosis (OR 10.84, 95% CI 5.76-20.40), cyclothymia (8.29, 4.65-14.76), schizoaffective disorder (7.90, 4.99-12.52), bipolar disorder (5.00, 4.23-5.90) and schizophrenia (4.37, 2.75-6.44); brief psychotic disorder, major depressive disorder, and anxiety disorders were also associated with elevated risk. Patterns were broadly similar in brother-sister pairs. Among women with no prior psychiatric diagnosis, sibling schizoaffective disorder (4.82, 2.99-7.74) and bipolar disorder (3.07, 2.59-3.64) were associated with particularly large relative-risk increases. Among women with a history of mania/psychosis, sibling psychiatric information contributed little additional predictive value.

**Interpretation:** Postpartum psychosis shows strong familial aggregation, with risks extending across multiple sibling diagnoses. The pattern of familial associations, with highest associations linked to sibling bipolar, cyclothymia, and schizoaffective disorder, supports conceptualizing postpartum psychosis within the spectrum of severe mood disorders with psychotic features.

## 1. Introduction

Postpartum psychosis is one of the most acute psychiatric emergencies associated with childbirth, affecting approximately 1-2 per 1,000 women ^1-3^. Onset typically occurs within weeks after delivery and is marked by rapid-onset mania, depression with psychotic features, mixed episodes, or psychotic symptoms without prominent mood symptomatology ^4^. Postpartum psychosis carries substantial risks for severe outcomes such as suicide, but has a good prognosis when detected in time, making early detection and intervention critical. Although a personal history of bipolar disorder remains the strongest known predictor, approximately half of affected women lack a prior diagnosis of severe mental illness ^5^, highlighting the need to identify additional risk factors.

Recent large-scale epidemiological studies have revealed that postpartum psychosis exhibits one of the strongest familial aggregation patterns among psychiatric disorders ^6^. In a nationwide Swedish cohort, having a full sister with postpartum psychosis conferred a 10.7-fold increased risk (95% CI, 6.6-16.3) of developing the condition ^6^. This relative recurrence risk exceeds that reported for many chronic psychiatric conditions, including bipolar disorder and schizophrenia ^7^. Postpartum psychosis is closely linked with bipolar disorder, given overlapping symptomatology, favorable treatment response to lithium and ECT, and a long-term course often marked by manic or depressive episodes ^5,8^. Polygenic risk score analyses indicate that women with postpartum psychosis share bipolar and schizophrenia risk alleles with women with bipolar disorder, but not major depression risk alleles, suggesting a partially distinct genetic architecture ^9,10^. Recent studies demonstrate that postpartum psychosis is highly heritable, with heritability of 55% based on familial data and 37% from genomic data ^9^. Previous Danish registry research found that family history of bipolar disorder increases risk of any postpartum psychiatric illness (HR = 2.86; 95% CI, 1.88-4.35), while family histories of schizophrenia and major depression confer more modest risks ^11^. However, these studies have not isolated postpartum psychosis specifically, nor focused on sibling diagnoses, a clinically accessible source of familial psychiatric risk.

Accordingly, in this population-based cohort study, we used Swedish national registers to evaluate whether having a full sibling with any psychiatric disorder is associated with increased risk of postpartum psychosis. We examined familial co-aggregation across a broad spectrum of psychiatric diagnoses, including mood, anxiety, psychotic, personality, neurodevelopmental, and substance use disorders. We additionally assessed whether familial associations differed by sibling sex and by the woman’s own psychiatric history, wherein the aforementioned diagnoses were grouped into one of the following categories: none, non-manic/non-psychotic, or manic/psychotic.

## 2. Methods

### 2.1 Study Population

This nationwide cohort study combined data from multiple Swedish national registers. Sweden assigns a unique personal identification number to all residents, enabling linkage across multiple national registers ^12^.

From the Swedish Medical Birth Register, we identified all women who delivered their first liveborn child between January 1, 1980, and October 31, 2017. This register includes more than 98% of births in Sweden ^13^. We required the index delivery to occur on or before October 31, 2017, to ensure a complete 90-day postpartum follow-up within the observation window.

Full siblings, defined as individuals sharing both biological parents, were identified using the Swedish Multi-Generation Register, which documents parent-child relationships with near-complete coverage (97% for mothers and 95% for fathers) (14). Women were included in the analytic cohort if at least one full sibling could be identified. Complete sibling ascertainment was achieved for individuals born in 1980 or later; see Sensitivity analyses for discussion of earlier birth cohorts.

The Swedish Ethical Review Authority approved the study and waived the requirement for informed consent owing to the use of de-identified registry data.

### 2.2 Exposure, Outcome, and Covariates

#### Diagnostic Sources

Diagnostic data were obtained from the Swedish National Patient Register, which includes all inpatient psychiatric care nationwide since 1987 and outpatient visits since 2001. Diagnoses are made by clinicians and registered using the International Classification of Diseases (ICD): ICD-8 (1969-1986), ICD-9 (1987-1996), and ICD-10 (1997-2017). The register has been extensively validated with positive predictive values for major psychiatric disorders, including bipolar disorder and schizophrenia-spectrum illnesses, exceeding 85% to 95%, and kappa statistics ranging from 0.74 to 0.87 for diagnostic agreement ^14,15^. Although specialist outpatient coverage was limited in earlier years and milder conditions may be underreported, postpartum psychosis generally requires hospitalization, thereby minimizing risk of outcome misclassification.

#### Outcome Definition

The primary outcome, postpartum psychosis, was defined as any psychiatric episode occurring within 90 days following a woman’s first liveborn delivery. Postpartum psychosis is one of the clearest phenotypes in psychiatry, but lacks accurate diagnostic criteria in major classification systems like the Diagnostic and Statistical Manual of Mental Disorders (DSM) ^16^. Therefore, to identify postpartum psychosis cases, we applied a diagnostic classification widely used in the literature: patients with mania and/or psychosis with onset in the postpartum period ^17^.

Similar to previous studies ^18^, case definition included postpartum-onset mania, psychosis, or depression with psychotic features, using the following diagnostic codes: ICD-10 (F23, F28, F29, F30, F31, F323, F333, F531); ICD-9 (296A, 296G, 298A, 298B, 298X); and ICD-8 (2961, 2963, 2968, 2969, 2981, 2944).

To maximize case ascertainment, we included both primary and secondary diagnoses with symptom onset within 0-3 months following a woman’s first liveborn childbirth. We selected this three-month period to ensure comprehensive coverage of the typical onset window and to account for delayed diagnosis, consistent with previous epidemiological studies ^3,4,19-21^.

#### Exposure Definition

The primary exposure for the index woman was having a full sibling with a psychiatric diagnosis recorded in the National Patient Register. We did not restrict exposure to older siblings; any full-sibling diagnosis was considered valid, and, for the risk estimation analyses, diagnoses were restricted to those occurring before the index woman’s first childbirth (anchor date) to ensure temporal ordering between exposure and outcome. For descriptive purposes, we additionally summarized lifetime sibling diagnoses, but all risk estimates were based on pre-childbirth exposures.

In the primary analysis, we included as covariates the calendar year of the index woman’s first delivery to account for temporal changes in diagnostic and reporting practices and the woman’s age at first childbirth to adjust for age-related risk differences.

### 2.3 Statistical Analysis

We first described cohort characteristics using medians and IQRs for continuous variables and counts and percentages for categorical variables.

To evaluate familial aggregation of postpartum psychosis, we fitted logistic regression models with postpartum psychosis as the binary outcome and sibling psychiatric diagnoses as the main exposures. Analyses were based on mother-sibling pairs identified in the pedigree, with each pair contributing a single observation. For each diagnostic category, we created binary indicators indicating whether the sibling had the diagnosis of interest, and we restricted exposures to diagnoses recorded before the woman’s first childbirth to preserve temporal ordering in the risk estimates. Psychotic-spectrum diagnoses, including delusional disorder, brief psychotic disorder, and psychosis not otherwise specified, were included in these diagnostic categories based on ICD-8, ICD-9 Swedish letter-code, and ICD-10 mappings. However, these exposures were extremely rare and yielded fewer than two postpartum psychosis cases among exposed sibling pairs. Consistent with our prespecified modeling rule to avoid unstable estimates, diagnostic categories with fewer than two outcome events were not analyzed further and therefore do not appear in the results tables. Cluster-robust standard errors were used in all models to account for non-independence of pairs within families. Models were adjusted for the woman’s age at first birth and calendar year of first birth. Familial associations were first estimated in sister-sister pairs and then in brother-sister pairs, and we also fitted combined models including both sisters and brothers to summarize overall sibling effects. Results are presented as odds ratios with 95% confidence intervals. All ICD-8, ICD-9 Swedish letter-code, and ICD-10 definitions used to construct each diagnostic category are provided in Supplementary Table S1.

To quantify the contribution of the woman’s own psychiatric history, we next examined associations between her pre-childbirth psychiatric diagnoses and postpartum psychosis in the full mother cohort. For each of the predefined diagnostic categories, we fitted logistic regression models with postpartum psychosis as the outcome and the presence of the diagnosis before the first birth as the exposure, adjusting for age and year at first childbirth.

We then evaluated whether sibling psychiatric history provided information beyond the woman’s own psychiatric background using hierarchical logistic regression. Mothers were classified into three mutually exclusive groups according to their pre-childbirth diagnoses: no prior psychiatric diagnosis; prior non-manic, non-psychotic disorders; and prior manic or psychotic disorders, defined as bipolar disorder, schizophrenia, or schizoaffective disorder. Within each stratum, we compared a base model that included maternal age and childbirth year, with or without an indicator of prior diagnosis, to an extended model that additionally included the sibling’s diagnosis of interest. Likelihood ratio tests were used to assess model improvement, and changes in Nagelkerke pseudo-R^2^ were calculated to quantify the incremental predictive value of sibling diagnoses. The odds ratio for the sibling diagnosis from the extended model was interpreted as the association between sibling history and postpartum psychosis within each level of the grouped pre-childbirth diagnoses.

Because multiple diagnostic categories were examined across families, we applied false discovery rate correction to p values from the sibling and maternal-history models using the Benjamini-Hochberg procedure, controlling the false discovery rate at 5%, to mitigate the risk of chance findings arising from multiple testing.

Sensitivity analyses assessed the robustness of key findings to birth cohort and sibship size. We repeated sibling analyses, adjusting for maternal birth cohort, and compared odds ratios from models with and without cohort adjustment. We also re-estimated bipolar sibling associations within categories of sibship size to examine whether estimates were consistent among women with one, two to three, or four or more siblings.

## 3. Results

### 3.1 Study Population and Descriptive Statistics

A total of 1,213,829 women with at least one full sibling were included in the analyses. Among these, 1,831 were diagnosed with postpartum psychosis within 90 days of their first liveborn delivery, corresponding to a prevalence of 1.5 per 1,000. The median maternal age at first delivery was 27 years (IQR 24-31), and the median year of childbirth was 1997 (IQR 1987-2008).

Among women with postpartum psychosis, 72.1% had at least one documented psychiatric diagnosis before childbirth, with just over half (53.2%) having bipolar disorder. Mood and anxiety disorders dominated the pre-childbirth psychiatric landscape: major depressive disorder affected more than one-third (36.3%), and anxiety disorders nearly one-third (29.3%).

Beyond mood disorders, substance use disorders (15.8%), brief psychotic disorder (14.9%), and personality disorders (14.3%) were also prevalent. Less common but clinically relevant diagnoses included stress/adjustment disorders (12.5%), ADHD (9.4%), and psychotic spectrum disorders such as schizophrenia (3.5%) and schizoaffective disorder (2.5%).

### 3.2 Familial Aggregation Analysis

The strongest familial association was postpartum psychosis itself: women whose sister had previously experienced postpartum psychosis had an approximately 11-fold increased risk (OR 10.84, 95% CI 5.76-20.40), representing the highest familial clustering observed across all psychiatric conditions examined.

Beyond this direct transmission, a broader pattern emerged linking a wide range of sibling psychiatric disorders to postpartum psychosis risk (Table 2). Disorders characterized by mood cyclicity showed the strongest associations. Cyclothymia conferred an approximately 8-fold increased risk (OR 8.29, 95% CI 4.65-14.76), while schizoaffective disorder showed nearly an 8-fold increase (OR 7.90, 95% CI 4.99-12.52). Bipolar disorder, the most prevalent serious psychiatric disorder among siblings, demonstrated an approximately 5-fold elevation in risk (OR 5.00, 95% CI 4.23-5.90).

**Table 1:** Distribution of Psychiatric Diagnoses before Childbirth Among Women with Postpartum Psychosis (N = 1,831)

| Diagnosis | Count | Percent |
| --- | --- | --- |
| Any psychiatric diagnosis | 1,320 | 72.1% |
| Bipolar disorder | 975 | 53.2% |
| Major depressive disorder | 665 | 36.3% |
| Anxiety disorders | 537 | 29.3% |
| Substance use disorders | 290 | 15.8% |
| Brief psychotic disorder | 272 | 14.9% |
| Personality disorders | 261 | 14.3% |
| Stress/adjustment disorders | 228 | 12.5% |
| ADHD | 172 | 9.4% |
| Eating disorders | 111 | 6.1% |
| Obsessive-compulsive disorder | 79 | 4.3% |
| Cyclothymia | 75 | 4.1% |
| Schizophrenia | 65 | 3.5% |
| Post-traumatic stress disorder | 64 | 3.5% |
| Sleep disorders | 51 | 2.8% |
| Schizoaffective disorder | 45 | 2.5% |
| Childhood behavioral disorders | 37 | 2.0% |
| Autism spectrum disorder | 36 | 2.0% |
| Delusional disorder | 29 | 1.6% |
| Intellectual disability | 26 | 1.4% |
| Dissociative disorders | 16 | 0.9% |
| Learning disorders | 8 | 0.4% |
| Tic disorders | 4 | 0.2% |

*Note: Percentages sum to more than 100% due to comorbidity among diagnoses.*

**Table 2.**
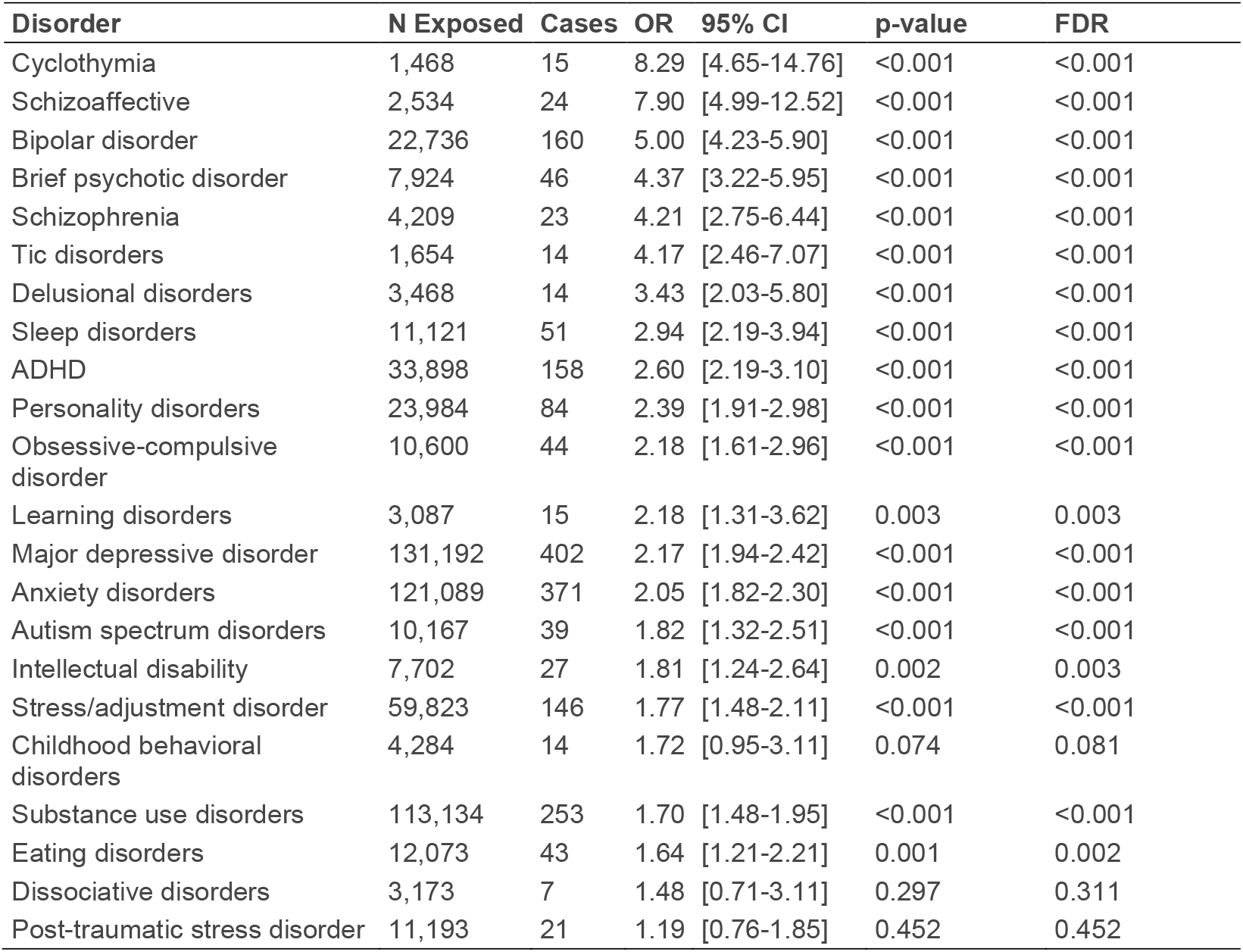
Combined Sibling Effects on Postpartum Psychosis.

| Disorder | N Exposed | Cases | OR | 95% CI | p-value | FDR |
| --- | --- | --- | --- | --- | --- | --- |
| Cyclothymia | 1,468 | 15 | 8.29 | [4.65-14.76] | <0.001 | <0.001 |
| Schizoaffective | 2,534 | 24 | 7.90 | [4.99-12.52] | <0.001 | <0.001 |
| Bipolar disorder | 22,736 | 160 | 5.00 | [4.23-5.90] | <0.001 | <0.001 |
| Brief psychotic disorder | 7,924 | 46 | 4.37 | [3.22-5.95] | <0.001 | <0.001 |
| Schizophrenia | 4,209 | 23 | 4.21 | [2.75-6.44] | <0.001 | <0.001 |
| Tic disorders | 1,654 | 14 | 4.17 | [2.46-7.07] | <0.001 | <0.001 |
| Delusional disorders | 3,468 | 14 | 3.43 | [2.03-5.80] | <0.001 | <0.001 |
| Sleep disorders | 11,121 | 51 | 2.94 | [2.19-3.94] | <0.001 | <0.001 |
| ADHD | 33,898 | 158 | 2.60 | [2.19-3.10] | <0.001 | <0.001 |
| Personality disorders | 23,984 | 84 | 2.39 | [1.91-2.98] | <0.001 | <0.001 |
| Obsessive-compulsive disorder | 10,600 | 44 | 2.18 | [1.61-2.96] | <0.001 | <0.001 |
| Learning disorders | 3,087 | 15 | 2.18 | [1.31-3.62] | 0.003 | 0.003 |
| Major depressive disorder | 131,192 | 402 | 2.17 | [1.94-2.42] | <0.001 | <0.001 |
| Anxiety disorders | 121,089 | 371 | 2.05 | [1.82-2.30] | <0.001 | <0.001 |
| Autism spectrum disorders | 10,167 | 39 | 1.82 | [1.32-2.51] | <0.001 | <0.001 |
| Intellectual disability | 7,702 | 27 | 1.81 | [1.24-2.64] | 0.002 | 0.003 |
| Stress/adjustment disorder | 59,823 | 146 | 1.77 | [1.48-2.11] | <0.001 | <0.001 |
| Childhood behavioral disorders | 4,284 | 14 | 1.72 | [0.95-3.11] | 0.074 | 0.081 |
| Substance use disorders | 113,134 | 253 | 1.70 | [1.48-1.95] | <0.001 | <0.001 |
| Eating disorders | 12,073 | 43 | 1.64 | [1.21-2.21] | 0.001 | 0.002 |
| Dissociative disorders | 3,173 | 7 | 1.48 | [0.71-3.11] | 0.297 | 0.311 |
| Post-traumatic stress disorder | 11,193 | 21 | 1.19 | [0.76-1.85] | 0.452 | 0.452 |

Other psychotic spectrum disorders beyond mood pathology also showed substantial associations: brief psychotic disorder (OR 4.37, 95% CI 3.22-5.95), schizophrenia (OR 4.21, 95% CI 2.75-6.44), and delusional disorder (OR 3.43, 95% CI 2.03-5.80) each at least tripled postpartum psychosis risk. More common psychiatric conditions, including major depressive disorder (OR 2.17, 95% CI 1.94-2.42) and anxiety disorders (OR 2.05, 95% CI 1.82-2.30), were associated with approximately 2-fold increases in risk.

Psychotic-spectrum conditions such as delusional disorder, brief psychotic disorder, and psychosis not otherwise specified were included in the diagnostic mappings but were omitted from some stratified analyses because they produced fewer than two postpartum psychosis cases among exposed sibling pairs, precluding stable estimation.

Sex-specific patterns emerged when examining sister-sister versus brother-sister concordance with postpartum psychosis (Figure 1; Supplementary Tables S2-S3). Psychotic spectrum disorders showed markedly stronger sister-to-sister risk: schizoaffective disorder conferred a 10-fold increase in risk through sisters (OR 10.44) versus 4-fold through brothers (OR 3.96), and brief psychotic disorder showed nearly 6-fold risk through sisters (OR 5.74) versus 2-fold through brothers (OR 2.42). Conversely, cyclothymia demonstrated stronger brother-to-sister concordance with postpartum psychosis (OR 12.67) compared with sister-to-sister (OR 7.44). For most other disorders, including bipolar disorder, depression, and anxiety, effect sizes remained broadly similar by sibling sex.

**Figure 1.**
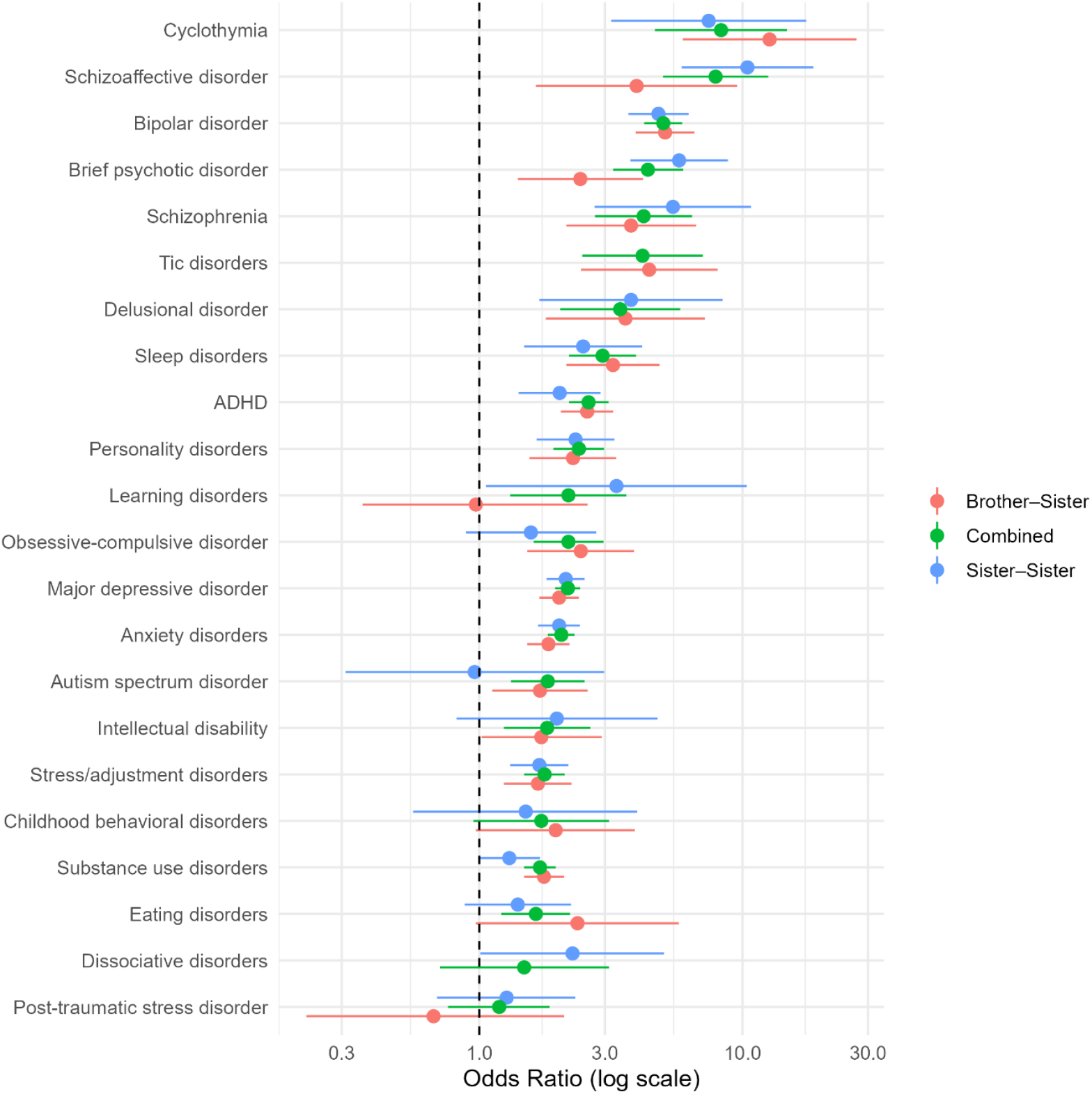
Familial Aggregation of Postpartum Psychosis

### 3.3 Independent Contribution of Sibling Psychiatric History

The predictive value of sibling psychiatric history varied by the woman’s own psychiatric status (Table 4). Among women with no prior psychiatric diagnosis, several sibling psychiatric disorders were strongly associated with postpartum psychosis. Schizoaffective disorder in a sibling was associated with nearly a five-fold increase in risk (OR 4.82, 95% CI 2.99-7.74), cyclothymia showed a similarly large association (OR 4.61, 95% CI 2.52-8.43), and bipolar disorder was associated with a three-fold increase in risk (OR 3.07, 95% CI 2.59-3.64). Brief psychotic episodes, schizophrenia, and several other disorders also substantially increased the risk in this psychiatrically healthy subgroup.

**Table 4.** Risk of postpartum psychosis associated with sibling psychiatric diagnoses after accounting for the woman’s own psychiatric history.

| Sibling Disorder | No psychiatric history<br>OR [95% CI] | Non-manic/psychotic<br>OR [95% CI] | Manic/psychotic<br>OR [95% CI] |
| --- | --- | --- | --- |
| ADHD | 1.43 [1.20-1.71] | 1.42 [0.90-2.23] | 1.07 [0.84-1.37] |
| Anxiety disorders | 1.26 [1.12-1.42] | 1.15 [0.81-1.64] | 1.10 [0.93-1.30] |
| Autism spectrum disorder | 1.07 [0.77-1.48] | - | 0.91 [0.60-1.37] |
| Bipolar disorder | 3.07 [2.59-3.64] | 1.46 [0.77-2.76] | 1.57 [1.23-2.01] |
| Brief psychotic disorder | 2.89 [2.12-3.96] | 1.60 [0.50-5.06] | 1.37 [0.82-2.28] |
| Childhood behavioral disorders | 0.69 [0.38-1.24] | - | 0.61 [0.30-1.27] |
| Cyclothymia | 4.61 [2.52-8.43] | - | 1.31 [0.42-4.06] |
| Delusional disorder | 2.02 [1.17-3.46] | - | 1.43 [0.57-3.61] |
| Major depressive disorder | 1.35 [1.21-1.51] | 1.02 [0.69-1.50] | 1.13 [0.97-1.32] |
| Dissociative disorders | 0.93 [0.44-1.97] | - | 0.58 [0.22-1.52] |
| Eating disorder | 1.15 [0.85-1.56] | 0.88 [0.33-2.39] | 0.77 [0.49-1.20] |
| Intellectual disability | 1.12 [0.76-1.64] | 0.95 [0.30-3.02] | 0.98 [0.59-1.65] |
| Learning disorders | 1.22 [0.73-2.05] | 1.25 [0.31-5.14] | 1.03 [0.53-2.00] |
| Obsessive-compulsive disorder | 1.33 [0.98-1.81] | 1.97 [0.90-4.32] | 1.01 [0.64-1.60] |
| Post-traumatic stress disorder | 0.64 [0.41-0.99] | 0.72 [0.23-2.23] | 0.57 [0.31-1.04] |
| Personality disorders | 1.28 [1.03-1.60] | 0.96 [0.48-1.93] | 1.03 [0.76-1.39] |
| Schizophrenia | 2.57 [1.67-3.95] | 1.93 [0.49-7.63] | 1.48 [0.80-2.71] |
| Schizoaffective disorder | 4.82 [2.99-7.74] | - | 2.09 [1.01-4.34] |
| Sleep disorders | 1.72 [1.28-2.32] | 0.83 [0.27-2.53] | 1.18 [0.79-1.74] |
| Stress reaction/<br>Adjustment disorders | 1.08 [0.90-1.28] | 0.96 [0.55-1.68] | 1.09 [0.85-1.39] |
| Substance use disorders | 1.03 [0.90-1.19] | 0.96 [0.61-1.51] | 0.88 [0.73-1.07] |
| Tic disorders | 2.41 [1.40-4.15] | 2.63 [0.65-10.54] | 1.36 [0.57-3.25] |

"-" denotes categories with insufficient exposed cases to generate reliable estimates; models did not converge or were not run for these strata.

In women with non-manic, non-psychotic psychiatric histories, the predictive value of sibling diagnoses was markedly reduced. Associations were generally attenuated and confidence intervals were wide, with no disorder showing a clear independent effect.

Among women with personal manic or psychotic histories, sibling psychiatric information contributed little additional predictive value. Most associations were close to the null; only bipolar disorder (OR 1.57, 95% CI 1.23-2.01) and schizoaffective disorder (OR 2.09, 95% CI 1.01-4.34) showed modest independent associations.

### 3.4 Sensitivity Analysis for Differential Sibling Ascertainment

Full numerical results from these sensitivity analyses are presented in Supplementary Tables S4-S6. Sensitivity analyses focused on the key sibling diagnoses that contributed the strongest and most stable associations in the primary models (bipolar disorder, schizoaffective disorder, schizophrenia, major depressive disorder, anxiety disorders, personality disorders, and substance use disorders). These conditions were selected because they account for the majority of familial aggregation observed in our main results and occurred with sufficient frequency to support sensitivity analyses.

Adjustment for maternal birth year had minimal impact on the familial associations. Although point estimates shifted slightly after cohort adjustment, in all cases the adjusted odds ratio lay within the 95% confidence interval of the corresponding unadjusted estimate, indicating that the magnitude of each association was largely unchanged. For example, the odds ratio for sibling schizophrenia changed from 3.69 (95% CI 2.45-5.57) to 4.21 (95% CI 2.79-6.36), while bipolar disorder (4.94 to 5.00), major depressive disorder (2.25 to 2.17), and anxiety disorders (2.29 to 2.05) showed similarly modest movements with substantial overlap in confidence intervals. The association with sibling schizoaffective disorder remained strong after adjustment (6.32 to 7.90), again well within the range of the unadjusted estimate.

Estimates were also similar across maternal birth cohorts: the sibling bipolar association remained elevated across women born in 1980-1989 (OR 3.35), 1990-1999 (OR 6.11), 2000-2009 (OR 4.82), and 2010 onward (OR 5.52). Finally, checks by sibship size indicated that the bipolar association was present across women with one sibling (OR 4.98), two to three siblings (OR 5.37), and four or more siblings (OR 3.57).

Overall, estimates for the main sibling diagnoses were similar after adjustment for maternal birth year and across maternal birth cohorts and sibship-size strata.

## 4. Discussion

In this nationwide cohort of more than 1.2 million Swedish women with at least one full sibling, we found strong familial aggregation of postpartum psychosis, with elevated risks associated with a broad range of sibling psychiatric diagnoses. The strongest association was for postpartum psychosis in a sister, which conferred an approximately eleven-fold increase in risk, followed by cyclothymia, schizoaffective disorder, and bipolar disorder. These findings extend prior work by demonstrating that familial liability for postpartum psychosis is not limited to bipolar or psychotic disorders but spans multiple diagnostic categories.

Consistent sex-specific patterns emerged. Across most diagnostic categories, associations were of similar magnitude in sister-sister and brother-sister pairs, but some psychotic-spectrum conditions, particularly schizoaffective disorder and brief psychotic disorder, showed stronger transmission through sisters than brothers. Although differences should be interpreted cautiously due to limited power for rarer disorders, these patterns suggest that both genetic and shared environmental factors contribute to familial clustering, with modest variation by sibling sex.

Stratification by the woman’s own psychiatric history revealed that sibling diagnoses are most informative among women with no prior psychiatric illness. In psychiatrically healthy women, sibling bipolar disorder, schizoaffective disorder, cyclothymia, schizophrenia, and brief psychotic disorder were all associated with substantial increases in postpartum psychosis risk. Among women with non-manic, non-psychotic disorders, associations were attenuated, and confidence intervals were wide. Among women with a history of manic or psychotic disorders, sibling diagnoses contributed little additional predictive value, indicating that personal psychiatric history dominates risk in this subgroup. Together, these findings highlight that family history may be particularly important for identifying elevated risk in clinically healthy women who lack standard risk markers.

Sensitivity analyses confirmed that the main results were robust to birth cohort effects and differences in sibship size. Adjusting for maternal birth cohort produced minimal changes in effect estimates, and sibling bipolar associations were consistent across women with small and large families. These findings reduce concern that incomplete sibling capture in older cohorts or variation in family size materially bias the observed associations.

Overall, our results strengthen the view that postpartum psychosis reflects a severe mood-psychotic vulnerability with a distinct familial signature. Although postpartum psychosis shares genetic and clinical features with bipolar disorder, the substantially stronger aggregation of postpartum psychosis in siblings than bipolar disorder suggests a familial liability that is partly specific to the postpartum phenotype. The broad pattern of cross-disorder aggregation, including associations with schizophrenia-spectrum disorders and mood-anxiety conditions, further indicates that multiple familial pathways may contribute to postpartum psychosis risk.

Several limitations merit consideration. First, sibling ascertainment is incomplete for women born before 1980, although sensitivity analyses indicate little impact on effect estimates. Second, use of registry-based diagnoses may under-capture milder psychiatric conditions or those treated exclusively in primary care. Third, some sibling-sex comparisons were limited by small sample sizes. Finally, the study design does not allow us to distinguish between genetic and shared environmental mechanisms underlying familial aggregation.

Future work integrating genomic data, hormonal markers, and detailed clinical phenotyping will be needed to clarify the biological pathways underlying familial risk for postpartum psychosis and to determine whether specific familial profiles predict treatment response or long-term outcomes. Identification of high-risk women through family history may ultimately support targeted monitoring and preventive strategies during the perinatal period.

## Data Availability Statement

Data may be obtained from a third party and are not publicly available. Data cannot be shared publicly owing to restrictions by law. Data are available from the National Medical Registries in Sweden after approval by the Swedish Ethical Review Authority.

## Acknowledgments

This study was supported by a grant from the Beatrice and Samuel A. Seaver Foundation (BM, SS); the National Institute of Mental Health (NIMH), R21MH131933 (BM), R01 HD111117 (TKR), and R01MH122869 (VB).

## Author Contributions

- Study concept and design: VB, BM, SS
- Acquisition, analysis, or interpretation of data: VB, BM, FM, TKR, SS
- Drafting of the manuscript: All authors
- Critical revision of the manuscript for important intellectual content: All authors
- Statistical analysis: BM, FM, SS
- Obtained funding: VB, BM
- Study supervision: VB, BM

## Notes

### Competing Interest Statement

The authors have declared no competing interest.

### Summary of Updates

Updated the source population and all the statistical analysis.

