## Supplementary Tables for "Sibling Psychiatric Disorders and Risk of Postpartum Psychosis"

**Table S1.** ICD-8, ICD-9 (Swedish), and ICD-10 Codes Used to Define Psychiatric Diagnostic Categories

| **Disorder** | **ICD-10** | **ICD-9 (Swedish)** | **ICD-8** |
| --- | --- | --- | --- |
| **Schizophrenia** | F20 | 295A, 295B, 295C, 295D, 295E, 295F, 295G, 295H, 295W, 295X | 29500, 29510, 29520, 29530, 29540, 29550, 29560, 29570, 29580, 29599 |
| **Schizoaffective disorder** | F25 | 295H | 29570 |
| **Delusional disorder** | F22 | 297B, 297C, 297D, 297W, 297X | 29700, 29710, 29798 |
| **Brief psychotic disorder** | F23 | 298B, 298C, 298E, 298W, 298X | 29800, 29810, 29820, 29830, 29899 |
| **Bipolar disorder** | F30, F31 | 296A, 296C, 296D, 296E, 296W, 298B | 29610, 29620, 29630, 29688, 29810 |
| **Major depressive disorder** | F32, F33, F341 | 296B, 298A, 300E, 300F, 311 | 29600, 29620, 29800, 30040 |
| **Cyclothymia** | F340 | 296W | 29630 |
| **Anxiety disorders** | F40, F41 | 300A, 300C | 30000, 30020 |
| **Obsessive–compulsive disorder** | F42 | 300D | 30030 |
| **Post-traumatic stress disorder** | F431 | 309B | — |
| **Stress reaction / Adjustment disorders** | F430, F432 | 308, 309A, 309X | 30799 |
| **ADHD** | F90 | 314J, 314W, 314X | 30899 |
| **Autism spectrum disorders** | F840, F841, F845, F849 | 299A, 299W, 299X | 29999 |
| **Intellectual disability** | F70–F79 | 317, 318A, 318B, 318C, 319 | 310–315 |
| **Tic disorders** | F95 | 307C | 30620 |
| **Learning disorders** | F81 | 315A, 315B, 315D, 315E, 315W, 315X | 30610 |
| **Eating disorders** | F50 | 307B, 307F | 30650 |
| **Substance use disorders** | F10–F19 | 291A–291X, 292A–292X, 303, 304, 305 | 29100, 29110, 29120, 29130, 29198, 29199, 29430, 303, 304 |
| **Personality disorders** | F60, F61 | 301A, 301B, 301C, 301D, 301E, 301F, 301G, 301H, 301J, 301W, 301X | 30100, 30110, 30120, 30130, 30140, 30150, 30160, 30170, 30180, 30188, 30199 |
| **Sleep disorders** | F51 | 307E | 30640 |
| **Dissociative disorders** | F44 | 300B | 30010 |
| **Childhood behavioural disorders** | F91, F93 | 312, 313A, 313B, 313C, 313D, 313W, 313X | 30899 |

**Table S2.** Sister-Sister Psychiatric Associations with Postpartum Psychosis

| **Psychiatric Disorder** | **N Exposed** | **Cases** | **OR (95% CI)** | **p-value** | **FDR** |
| --- | --- | --- | --- | --- | --- |
| Schizoaffective disorder | 1,476 | 16 | 10.44 (5.87–18.59) | <0.001 | <0.001 |
| Cyclothymia | 939 | 8 | 7.44 (3.17–17.46) | <0.001 | <0.001 |
| Brief psychotic disorder | 3,898 | 25 | 5.74 (3.75–8.80) | <0.001 | <0.001 |
| Schizophrenia | 1,771 | 10 | 5.44 (2.74–10.77) | <0.001 | <0.001 |
| Bipolar disorder | 12,181 | 72 | 4.79 (3.68–6.24) | <0.001 | <0.001 |
| Delusional disorder | 1,537 | 6 | 3.77 (1.69–8.41) | 0.001 | 0.002 |
| Learning disorders | 500 | 3 | 3.32 (1.06–10.38) | 0.039 | 0.061 |
| Sleep disorders | 4,909 | 16 | 2.48 (1.48–4.16) | <0.001 | 0.001 |
| Personality disorders | 12,260 | 38 | 2.32 (1.65–3.26) | <0.001 | <0.001 |
| Dissociative disorders | 1,977 | 6 | 2.26 (1.01–5.03) | 0.046 | 0.069 |
| Major depressive disorder | 68,512 | 180 | 2.13 (1.80–2.51) | <0.001 | <0.001 |
| ADHD | 10,485 | 32 | 2.02 (1.41–2.89) | <0.001 | <0.001 |
| Anxiety disorders | 61,948 | 161 | 2.01 (1.67–2.41) | <0.001 | <0.001 |
| Intellectual disability | 1,711 | 5 | 1.97 (0.82–4.76) | 0.130 | 0.161 |
| Stress/adjustment disorders | 32,311 | 71 | 1.69 (1.31–2.18) | <0.001 | <0.001 |
| Obsessive-compulsive disorder | 4,995 | 12 | 1.57 (0.89–2.78) | 0.118 | 0.155 |
| Childhood behavioral disorders | 1,658 | 4 | 1.50 (0.56–3.98) | 0.419 | 0.440 |
| Eating disorders | 7,370 | 18 | 1.40 (0.88–2.23) | 0.160 | 0.192 |
| Substance use disorders | 37,653 | 62 | 1.30 (0.99–1.70) | 0.055 | 0.077 |
| Post-traumatic stress disorder | 6,715 | 12 | 1.27 (0.69–2.32) | 0.442 | 0.453 |
| Autism spectrum disorder | 1,994 | 3 | 0.96 (0.31–2.98) | 0.944 | 0.944 |

Note: Odds ratios adjusted for maternal age at first birth and year of first birth, with cluster-robust standard errors to account for family clustering.

**Table S3.** Brother-Sister Psychiatric Associations with Postpartum Psychosis

| **Psychiatric Disorder** | **N Exposed** | **Cases** | **OR (95% CI)** | **p-value** | **FDR** |
| --- | --- | --- | --- | --- | --- |
| Cyclothymia | 441 | 7 | 12.67 (5.93–27.10) | <0.001 | <0.001 |
| Bipolar disorder | 8,462 | 59 | 5.08 (3.92–6.57) | <0.001 | <0.001 |
| Tic disorders | 1,181 | 11 | 4.42 (2.43–8.04) | <0.001 | <0.001 |
| Schizoaffective disorder | 910 | 5 | 3.96 (1.64–9.55) | 0.002 | 0.003 |
| Schizophrenia | 2,218 | 12 | 3.77 (2.14–6.66) | <0.001 | <0.001 |
| Delusional disorder | 1,794 | 8 | 3.59 (1.79–7.20) | <0.001 | <0.001 |
| Sleep disorders | 5,009 | 25 | 3.22 (2.14–4.84) | <0.001 | <0.001 |
| ADHD | 18,911 | 88 | 2.57 (2.04–3.22) | <0.001 | <0.001 |
| Obsessive-compulsive disorder | 3,786 | 18 | 2.43 (1.52–3.87) | <0.001 | <0.001 |
| Brief psychotic disorder | 3,619 | 13 | 2.42 (1.40–4.19) | 0.002 | 0.002 |
| Eating disorders | 950 | 5 | 2.36 (0.97–5.73) | 0.057 | 0.065 |
| Personality disorders | 8,777 | 27 | 2.27 (1.55–3.31) | <0.001 | <0.001 |
| Major depressive disorder | 50,681 | 142 | 2.01 (1.69–2.39) | <0.001 | <0.001 |
| Childhood behavioral disorders | 2,055 | 8 | 1.95 (0.97–3.90) | 0.060 | 0.065 |
| Anxiety disorders | 45,858 | 126 | 1.83 (1.52–2.20) | <0.001 | <0.001 |
| Substance use disorders | 68,618 | 152 | 1.76 (1.48–2.10) | <0.001 | <0.001 |
| Intellectual disability | 4,036 | 14 | 1.72 (1.02–2.92) | 0.043 | 0.051 |
| Autism spectrum disorder | 5,956 | 22 | 1.70 (1.12–2.58) | 0.013 | 0.017 |
| Stress/adjustment disorders | 23,553 | 52 | 1.67 (1.24–2.24) | <0.001 | 0.001 |
| Learning disorders | 1,821 | 4 | 0.97 (0.36–2.58) | 0.944 | 0.944 |
| Post-traumatic stress disorder | 2,933 | 3 | 0.67 (0.22–2.10) | 0.495 | 0.521 |
| Dissociative disorders | — | — | — | — | — |

Note: Odds ratios adjusted for maternal age at first birth and year of first birth, with cluster-robust standard errors to account for family clustering.

**Table S4.** Impact of Maternal Birth Year Adjustment on Sibling Associations

| **Psychiatric Disorder** | **N Exposed** | **Cases** | **OR Without Birth Year Adjustment (95% CI)** | **OR With Birth Year Adjustment (95% CI)** | **% Change** |
| --- | --- | --- | --- | --- | --- |
| Schizoaffective disorder | 2,534 | 24 | 6.32 (4.22–9.47) | 7.90 (5.27–11.85) | +24.99% |
| Schizophrenia | 4,209 | 23 | 3.69 (2.45–5.57) | 4.21 (2.79–6.36) | +13.97% |
| Substance use disorders | 113,134 | 253 | 1.61 (1.42–1.83) | 1.70 (1.49–1.93) | +5.45% |
| Bipolar disorder | 22,736 | 160 | 4.94 (4.21–5.79) | 5.00 (4.26–5.87) | +1.31% |
| Major depressive disorder | 131,192 | 402 | 2.25 (2.03–2.50) | 2.17 (1.95–2.41) | –3.89% |
| Personality disorders | 23,984 | 84 | 2.54 (2.05–3.16) | 2.39 (1.92–2.97) | –6.14% |
| Anxiety disorders | 121,089 | 371 | 2.29 (2.05–2.55) | 2.05 (1.83–2.28) | –10.64% |

Note: Both models use cluster-robust standard errors. Birth year adjustment includes maternal year of birth as a covariate.

**Table S5.** Sibling Bipolar Disorder Association by Maternal Birth Cohort

| **Birth Cohort** | **N Mothers** | **N Exposed to Sibling Bipolar** | **PPS Cases** | **OR (95% CI)** |
| --- | --- | --- | --- | --- |
| 1980–1989 | 704,890 | 8,027 | 18 | 3.35 (2.09–5.37) |
| 1990–1999 | 441,421 | 5,133 | 24 | 6.11 (4.04–9.26) |
| 2000–2009 | 491,779 | 5,703 | 36 | 4.82 (3.44–6.74) |
| 2010–2017 | 370,730 | 3,873 | 82 | 5.52 (4.41–6.91) |

Note: Odds ratios adjusted for maternal age at first birth, with cluster-robust standard errors.

**Table S6.** Sibling Bipolar Disorder Association by Number of Siblings

| **Sibship Size** | **N Mother-Sibling Pairs** | **OR (95% CI)** |
| --- | --- | --- |
| 1 sibling | 685,090 | 4.98 [3.73-6.64] |
| 2–3 siblings | 1,057,228 | 5.37 [4.29-6.72] |
| 4+ siblings | 266,502 | 3.57 [2.26-5.62] |

Note: Odds ratios adjusted for maternal age and year at first birth, with cluster-robust standard errors.
